# Safety and Immunogenicity of Inactivated SARS-CoV-2 Vaccine in High-Risk Occupational Population: a randomized, parallel, controlled clinical trial

**DOI:** 10.1101/2021.08.06.21261696

**Authors:** Yongliang Feng, Jing Chen, Tian Yao, Yue Chang, Xiaoqing Li, Rongqin Xing, Hong Li, Ruixue Xie, Xiaohong Zhang, Zhiyun Wei, Shengcai Mu, Ling Liu, Lizhong Feng, Suping Wang

**Author notes:** Author for correspondence: Suping Wang, professor,; Address: Department of Epidemiology, School of Public Health, Shanxi Medical University, 56 Xinjian South Road Taiyuan, 030001, Shanxi Province, China, Lizhong Feng, chief physician,; Address: Shanxi Provincial Center for Disease Control and Prevention, 8 Xiaonanguan Street Taiyuan, 030012, Shanxi Province, China. Co-first authors (Yongliang Feng, Jing Chen, Tian Yao).

## Abstract

Vaccination is urgently needed to prevent the global spread of severe acute respiratory syndrome coronavirus 2 (SARS-CoV-2). Here, we conducted a randomized, parallel, controlled clinical trial for assessment of the immunogenicity and safety of an inactivated SARS-CoV-2 vaccine, aiming to determine an appropriate vaccination interval for high-risk occupational population. Participants were randomly assigned to receive two doses of inactivated SARS-CoV-2 vaccine (4 µg per dose) at an interval of either 14 days, 21 days or 28 days. The primary immunogenicity endpoints were neutralization antibody seroconversion and geometric mean titer (GMT) at 28 days after the second dose. Our results showed that the seroconversion rates (GMT ≥ 16) were all 100% in the three groups and the 0-21 and 0-28 groups elicited significantly higher SARS-CoV-2 neutralizing antibody level. All reported adverse reactions were mild. (Chinese Clinical Trial Registry: ChiCTR2100041705, ChiCTR2100041706)

## Introduction

The ongoing pandemic of coronavirus disease 2019 (COVID-19) induced by severe acute respiratory syndrome coronavirus 2 (SARS-CoV-2) has led to an unprecedented global public health crisis. Globally, as of August 4, 2021, more than 199 million cases of SARS-CoV-2 infection and more than 4.2 million deaths have been reported^1^. SARS-CoV-2 belongs to the Betacoronavirus of the family Coronaviridae, and commonly induces a spectrum of clinical manifestations ranging from asymptomatic, minor flu-like symptoms to acute respiratory distress syndrome (ARDS), pneumonia and even death^2^. Compared with other coronaviruses, SARS-CoV-2 appears to undergo more rapid transmission and variation^3, 4^. Although it is proved to be effective that the COVID-19 pandemic can be controlled using strict social hygiene measures such as physical distancing and masks, the absence of herd immunity leaving people susceptible to further waves of SARS-CoV-2 infection, especially for the high-risk occupational population. Meantime, the measures taken to contain SARS-CoV-2 have placed a substantial burden on health-care systems around the world, with far-reaching social and economic consequences. Hence, a safe and effective vaccine against COVID-19 is urgently needed to prevent the resurgence of the epidemic.

Many countries have accelerated the process of clinical trials to determine an effective and safe vaccine to prevent COVID-19 pandemic. Currently, more than 292 candidate vaccines are in development worldwide, 37 of which are already in phase 3 trials using different platforms^5^, including nucleic acid (DNA and RNA) vaccines, viral vector (replicating and non-replicating) vaccines, virus-like particles vaccines, peptide-based vaccines, recombinant protein vaccines and inactivated vaccines^6-8^. Inactivated vaccines have been widely used against various infectious diseases for decades. Their long history of use confers some advantages, such as well-developed and mature manufacturing processes, ease of scaling up production and storage, and the ability to present multiple viral proteins for immune recognition. In addition, inactivated vaccines induce high levels of neutralizing antibody titers in mice, rats, guinea pigs, rabbits, and nonhuman primates to provide protection against SARS-CoV-2^9-11^. Moreover, the results of previous clinical trials on the inactivated vaccines conducted in several countries showed good neutralizing antibody responses and/or efficacy against disease caused by COVID-19^12-21^. To date, two inactivated SARS-CoV-2 vaccines manufactured by the Beijing Institute of Biological Products/Sinopharm (China) and Sinovac Life Sciences/CoronaVac (China) have been placed on WHO’s Emergency Use Listing. Here, we report the analysis of immunogenicity and safety of an inactivated SARS-CoV-2 vaccine manufactured by Beijing Institute of Biological Products Co., Ltd (China).

Previous studies^15-17, 22-24^ have shown that the three immunization programs (0, 14 procedure, 0, 21 procedure or 0, 28 procedure) induce varying degrees of immune effect, but the optimal interval of injections remains unclear. Furthermore, the safety and immunogenicity of inactivated SARS-CoV-2 vaccine in occupational high-risk population have not been reported. Therefore, based on the preliminary clinical trials, we explored the immunogenicity and safety of the three different SARS-CoV-2 inactivated vaccination schemes at an interval of either 14 days, 21 days or 28 days in high-risk occupational population to optimize the inactivated vaccination regimen. We would continue to follow up until months 3, 6, and 12 in the further study.

## Methods

### Study design and participants

We conducted a randomized, controlled clinical trial of the SARS-CoV-2 inactivated vaccine manufactured by Beijing Biological Products Institute Co., Ltd. in Taiyuan City, Shanxi Province, China. Written informed consents were obtained from all participants before enrollment. Eligible participants were people aged 18-59 years, signed the informed consent form and participated voluntarily with good compliance. Exclusion criteria were participants with history or family history of allergy, convulsion, epilepsy, encephalopathy or psychosis; any intolerance or allergy to any component of the vaccine; known or suspected diseases including severe respiratory disease, severe cardiovascular disease, severe liver or kidney disease, medically uncontrollable hypertension (systolic blood pressure ≥ 140 mmHg and diastolic blood pressure ≥ 90 mmHg), complications of diabetes mellitus, malignancy, various acute diseases or acute episodes of chronic disease; various infectious, suppurative and allergic skin diseases; congenital or acquired immunodeficiency, other vaccination history within 14 days before vaccination, a history of coagulation dysfunction, a history of non-specific immunoglobulin injection within 1 month prior to enrollment, acute illness with fever (body temperature > 37.0°C); and being pregnant or breastfeeding.

The protocol was approved by the Ethics Committee of Shanxi Provincial Center for Disease Control and Prevention and was conducted in accordance with the Declaration of Helsinki and Good Clinical Practice. All participants signed a consent form after being informed about the study. The trial was registered with ChiCTR.org.cn (ChiCTR2100041705, ChiCTR2100041706).

### Procedures

A computerized random number generator performed block randomization with a randomly selected block size of 6, and eligible participants were randomly assigned into three groups to receive two doses inactivated COVID-19 vaccine at the schedule of day 0-14, day 0-21, or day 0-28. Each dose of vaccine containing 4 µg of inactivated SARS-CoV-2 virus antigen was intramuscularly injected into the lateral deltoid muscle of the upper arm. The vaccines used in this study were inactivated vaccine (Vero Cell) produced by Beijing Biological Products Institute Co., Ltd. Demographic information (age, gender, body mass index (BMI), marital status, and education level), influenza vaccination history, smoking, drinking, and chronic diseases were collected via questionnaire investigation.

### Safety assessment

After each dose was vaccinated, the participants were observed for any immediate reaction for 30 min, and local and systemic adverse reactions were collected. Participants were required to record the local adverse events and systemic adverse events on diary cards within 7 days of each injection. Any other unsolicited symptoms were also recorded during a 28-day follow-up period after each injection by spontaneous report from the participants combined with the regular visit. The solicited adverse reactions included local reactions (pain, induration, swelling, rash, flush, and pruritus) and systematic reactions (fever, diarrhea, dysphagia, anorexia, vomiting, nausea, muscle pain (non-vaccination sites), arthralgia, headache, cough, dyspnea, skin and mucosal abnormalities, acute allergic reactions, and fatigue).

### Laboratory methods

Oropharyngeal/nasal swabs were collected for detecting SARS-CoV-2 nucleic acid from all subjects by using reverse transcriptase-polymerase chain reaction (RT-PCR) test before the first dose of vaccine vaccination, before the second dose of vaccine vaccination, 28 days after the whole course of vaccination, respectively. Blood samples were taken from participants for serology tests before the first injection and on day 28 after the second injection. The neutralizing antibody to live SARS-CoV-2 (strain 19nCoV-CDC-Tan-Strain 05 [QD01]) were quantified using a micro cytopathogenic effect assay at baseline and 28 days after immunization. We defined the neutralizing antibody seroconversion rate as post-injection titer of a 16-fold. (Baseline titers were all below 1:4).

### Outcomes

The primary immunogenic endpoints were the seroconversion rates (geometric mean titer (GMT) ≥ 16) and GMT of neutralizing antibody to live SARS-CoV-2 at day 28 after the last dose. Secondary immunogenic endpoints were the positive rates (GMT ≥ 32, 64, 128, 256) and GMT of neutralizing antibody to live SARS-CoV-2, 28 days after the whole course of vaccination, respectively. The primary endpoint for safety was the occurrence of adverse reactions within 7 days after the first and second vaccinations. Adverse events within 28 days after the first and the second vaccinations across the three groups were analyzed as secondary safety endpoints.

### Statistical analysis

The study sample size of 360 participants provided 84.4% power to detect a difference of 5% (85% vs. 80%) of responders in the 0-21 and 0-28 vaccination groups compared with the 0-14 group, respectively in airport ground staff and public security officers. Data were recorded using EpiData, and analyses were performed using SAS version 9.3 (SAS Institute, Cary, NC, USA). Analysis of variance (ANOVA) was used to analyze continuous data, and the chi-square or Fisher’s exact test was used for categorical data. We assessed immunogenic endpoints by the modified intention-to-treat (ITT) analysis (ie, subjects who undertake randomization) and per-protocol (PP) analysis (ie, subjects who compliant to the protocol, receive 2 doses of vaccine according to the requirements of the protocol, and have serum-testing results before and after immunization). Multinomial logistic regression analysis and unconditional logistic regression model was used to determine the influencing factors of SARS-CoV-2 neutralizing antibody immunization. The safety analysis was performed on data from all subjects who received vaccination after randomization. The level of statistical significance for all analyses was *P* < 0.05.

## Results

### Study participants and baseline characteristics

Between January and May 2021, 810 participants were screened, and 809 were enrolled (73.18% male, 26.82% female; mean age 38.78 years). Of the 809 participants who were enrolled, 405 participants were the public security officers and 404 participants were the airport ground staff. 270 participants were included in the group 0-14 vaccination cohort, 270 participants in the group 0-21 vaccination cohort, and 269 participants in the group 0-28 vaccination cohort. Among them, 809 (100%) patients received the first injection and completed the two-dose vaccination schedule, with 270, 270, and 269 patients in the 0-14, 0-21, and 0-28 groups, respectively. A total of 256, 247 and 241 patients in the 0-14, 0-21, and 0-28 groups, respectively, completed the follow-up 28 days after the whole course of vaccination (Figure 1). The baseline characteristics of the participants are shown in Table 1. There were no significant differences in demographic and behavioral characteristics among the three groups at baseline and 28 days after the whole course of vaccination (*P* > 0.05; Table 1, Supplement 1).

**Table 1.** Baseline Characteristics of High-risk Occupational Population with Different Vaccinations.

| Characteristics | Total (n=809) | 0-14 group (n=270) | 0-21 group (n=270) | 0-28 group(n=269) | *P* |
| --- | --- | --- | --- | --- | --- |
| Gender |  |  |  |  | 0.449 |
| Male | 592(73.18) | 192(71.11) | 196(72.59) | 204(75.84) |  |
| Female | 217(26.82) | 78(28.89) | 74(27.41) | 65(24.16) |  |
| Age (year) |  |  |  |  | 0.229 |
| <40 | 463(57.23) | 144(53.33) | 156(57.78) | 163(60.59) |  |
| 40~ | 346(42.77) | 126(46.67) | 114(42.22) | 106(39.41) |  |
| Education level |  |  |  |  | 0.335 |
| Junior high school or lower | 74(9.15) | 31(11.48) | 25(9.26) | 18(6.69) |  |
| Senior high school | 37(4.57) | 10(3.70) | 12(4.44) | 15(5.58) |  |
| College or higher | 698(86.28) | 229(84.82) | 233(86.30) | 236(87.73) |  |
| Ethnicity |  |  |  |  | 0.366<sup>a</sup> |
| Han ethnicity | 797(98.52) | 268(99.26) | 264(97.78) | 265(98.51) |  |
| Other | 12(1.48) | 2(0.74) | 6(2.22) | 4(1.49) |  |
| Marital status |  |  |  |  | 0.267 |
| Married | 621(76.76) | 217(80.37) | 196(72.59) | 208(77.32) |  |
| Unmarried | 165(20.40) | 47(17.41) | 66(24.45) | 52(19.33) |  |
| Divorced or widowed | 23(2.84) | 6(2.22) | 8(2.96) | 9(3.35) |  |
| BMI (kg/m<sup>2</sup>) |  |  |  |  | 0.848 |
| <18.5 | 19(2.35) | 8(2.96) | 6(2.22) | 5(1.86) |  |
| 18.5~ | 333(41.16) | 110(40.74) | 116(42.97) | 107(39.78) |  |
| 24~ | 457(56.49) | 152(56.30) | 148(54.81) | 157(58.36) |  |
| Influenza vaccination history |  |  |  |  | 0.865 |
| No | 549(67.86) | 180(66.67) | 184(68.15) | 185(68.77) |  |
| Yes | 260(32.14) | 90(33.33) | 86(31.85) | 84(31.23) |  |
| Occupation |  |  |  |  | 0.991 |
| Public security officers | 405(50.06) | 135(50.00) | 136(50.37) | 134(49.81) |  |
| Airport ground staff | 404(49.94) | 135(50.00) | 134(49.63) | 135(50.19) |  |
| Smoking |  |  |  |  | 0.368 |
| No | 545(67.37) | 190(70.37) | 181(67.04) | 174(64.68) |  |
| Yes | 264(32.63) | 80(29.63) | 89(32.96) | 95(35.32) |  |
| Drinking |  |  |  |  | 0.906 |
| No | 621(76.76) | 206(76.30) | 206(76.30) | 209(77.70) |  |
| Yes | 188(23.24) | 64(23.70) | 64(23.70) | 60(22.30) |  |
| Chronic diseases |  |  |  |  | 0.978 |
| No | 754(93.20) | 252(93.33) | 252(93.33) | 250(92.94) |  |
| Yes | 55(6.80) | 18(6.67) | 18(6.67) | 19(7.06) |  |

Results expressed as n (%); <sup>a</sup> Fisher's exact test.

**Figure 1.**
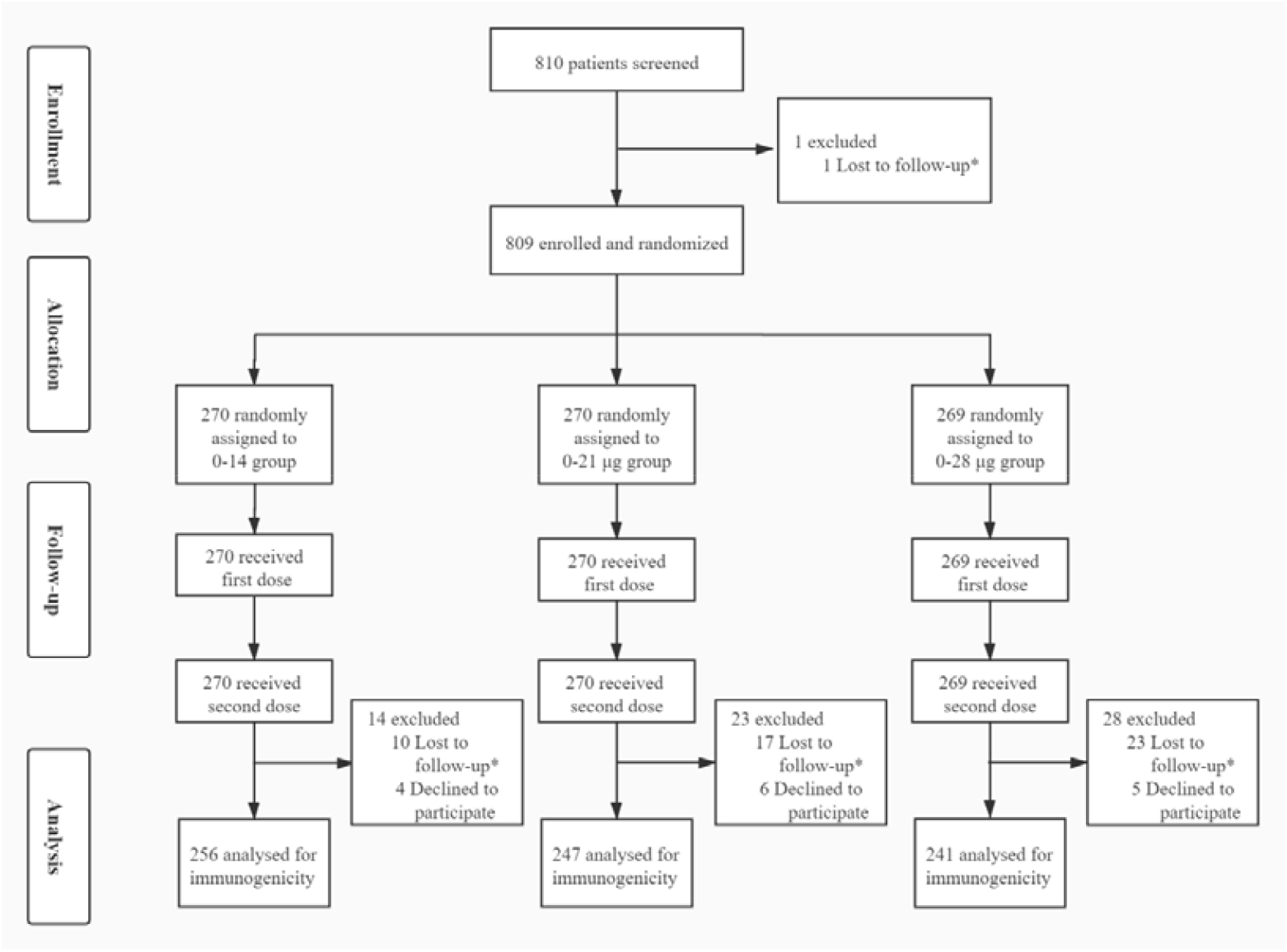
Flow of Participants in a Study of the Inactivated SARS-CoV-2 Vaccine in high-risk occupational population. * Lost to follow up including not being at the study site, or illness.

### Assessment of immunity elicited by the vaccine in the three immunization procedures

The seroconversion rate and GMT of SARS-CoV-2 neutralizing antibody in the three groups By day 28 after the second injection, the seroconversion rates of neutralizing antibody (GMT ≥ 16) were all 100% in the 0-14, 0-21, and 0-28 groups. SARS-CoV-2 neutralizing antibody with a GMT of 98.41 (95% *CI*, 88.38-108.40) was noted in the 0-14 group, which was significantly lower compared with 134.40 (95% *CI*, 123.10-145.70) in the 0-21 group (*P* < 0.001 vs. 0-14 group) and 145.50 (95% *CI*, 131.30-159.60) in the 0-28 group (*P* < 0.001 vs. 0-14 group). Moreover, the GMT of neutralizing antibody in 0-21 group was numerically higher than that in 0-28 group (*P* = 0.228) (Table 2). The modified ITT analysis showed that the seroconversion rates of neutralizing antibody were 94.81% (256/270) in the 0-14 group, 91.48% (247/270) in the 0-21 group (*P* > 0.05 vs. 0-14 group) and 89.59% (241/269) in the 0-28 group (*P* > 0.05 vs. 0-14 group), with the neutralizing antibody GMT of 93.51 (95% *CI*, 83.68-103.30), 122.00 (95% *CI*, 110.70-133.20), and 129.80 (95% *CI*, 116.20-143.40) in the three groups, respectively (Table 2).

**Table 2.**
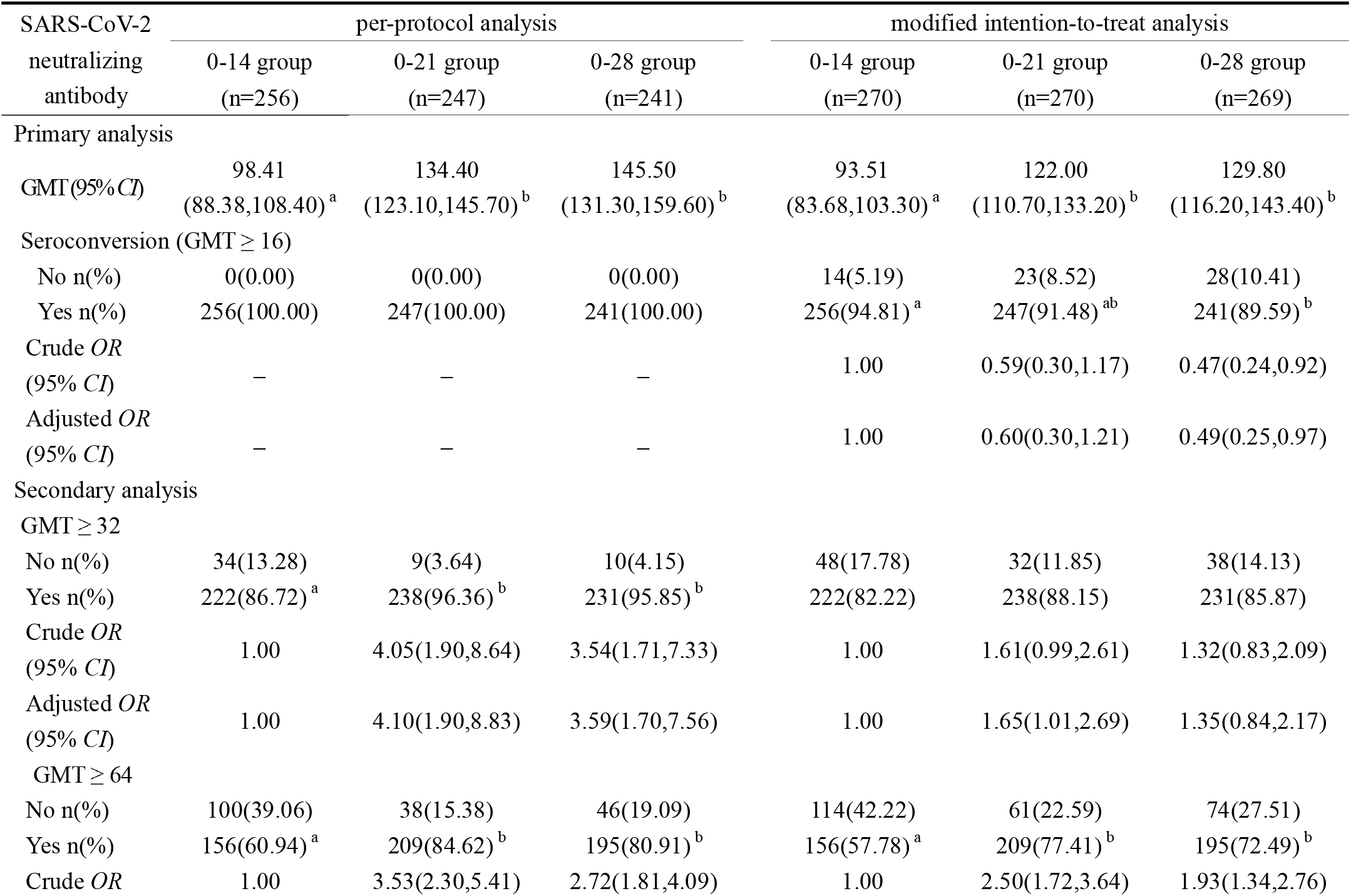
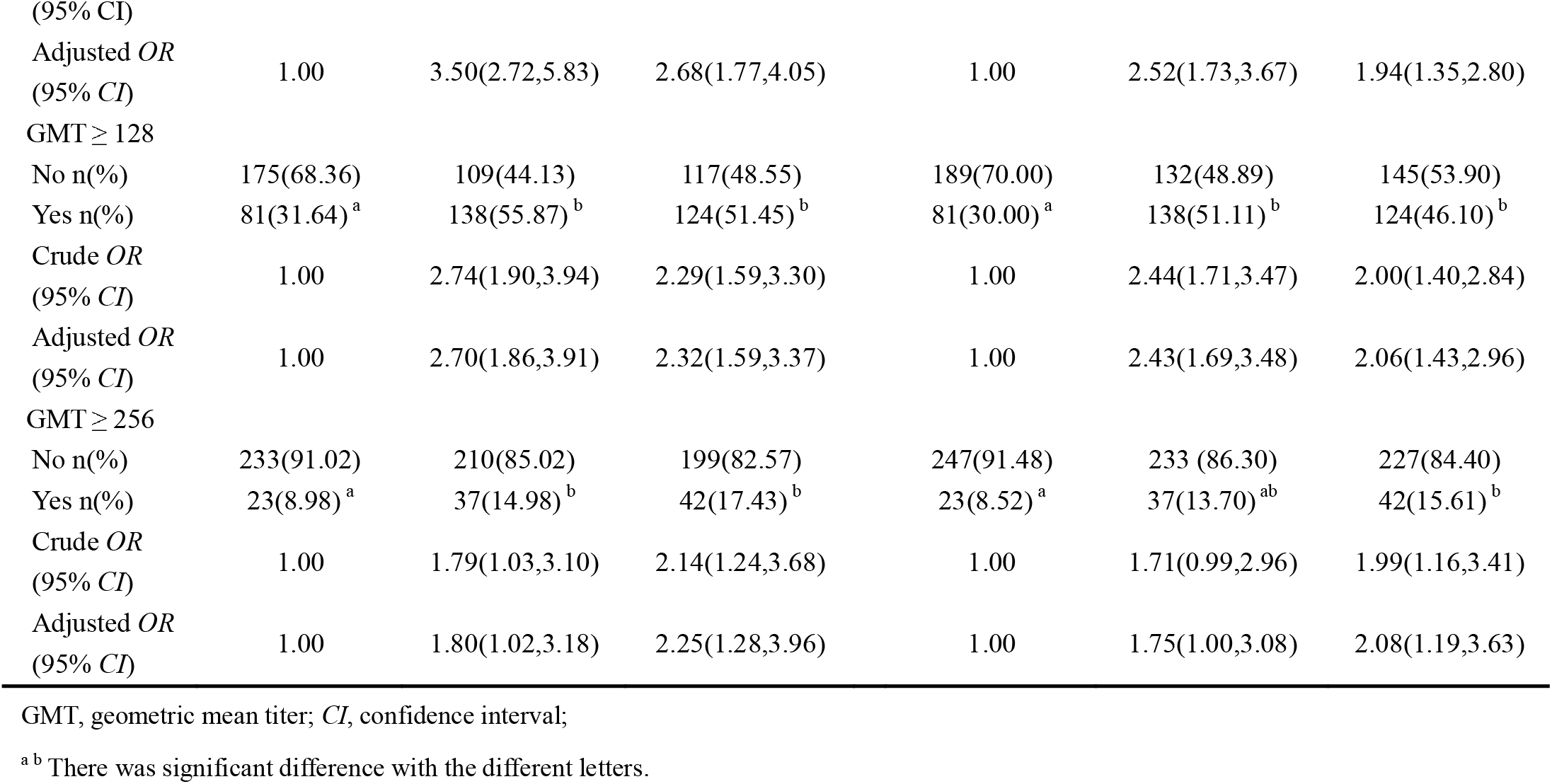
The seroconversion rate and GMT of SARS-CoV-2 Neutralizing antibody in the three groups.

Then we used different criteria to determine the immunization of neutralizing antibody for comparison. The positive rates of neutralizing antibody GMT ≥ 32 were 86.72% (222/256), 96.36% (238/247; *P* < 0.001) and 95.85% (231/241; *P* < 0.001) in the 0-14, 0-21, and 0-28 groups, respectively. The positive rates of neutralizing antibody GMT ≥ 64, 128, and 256 were 60.94% (156/256), 31.64% (81/256) and 8.98% (23/256) in the 0-14 group, 84.62% (209/247), 55.87% (138/247) and 14.98% (37/247) in the 0-21 group, 80.91% (195/241), 51.45% (124/241) and 17.43% (42/241) in the 0-28 group, respectively (Table 2). The positive rates of neutralizing antibody (GMT ≥ 32, 64, 128, or 256) in the 0-21 and 0-28 groups were significantly higher than that in the 0-14 group, and there was no significant difference between the 0-21 group and 0-28 group. The modified ITT analysis yielded similar results (Table 2).

#### Distribution of SARS-CoV-2 neutralizing antibody in the three groups

In the 0-14 group, the proportion of neutralizing antibody was higher at the GMT 32-63 (25.78%, 66/256) and 64-127 (29.30%, 75/256), while the proportion of neutralizing antibody in the 0-21 and 0-28 groups was higher at the GMT of 64-127 (28.74%, 71/247; 29.46%, 71/241) and 128-255 (40.90%, 101/247; 34.02%, 82/241), respectively. (Figure 2). There were significant differences in distribution of SARS-CoV-2 neutralizing antibody GMT among the three vaccination groups (*P* < 0.001).

**Figure 2.**
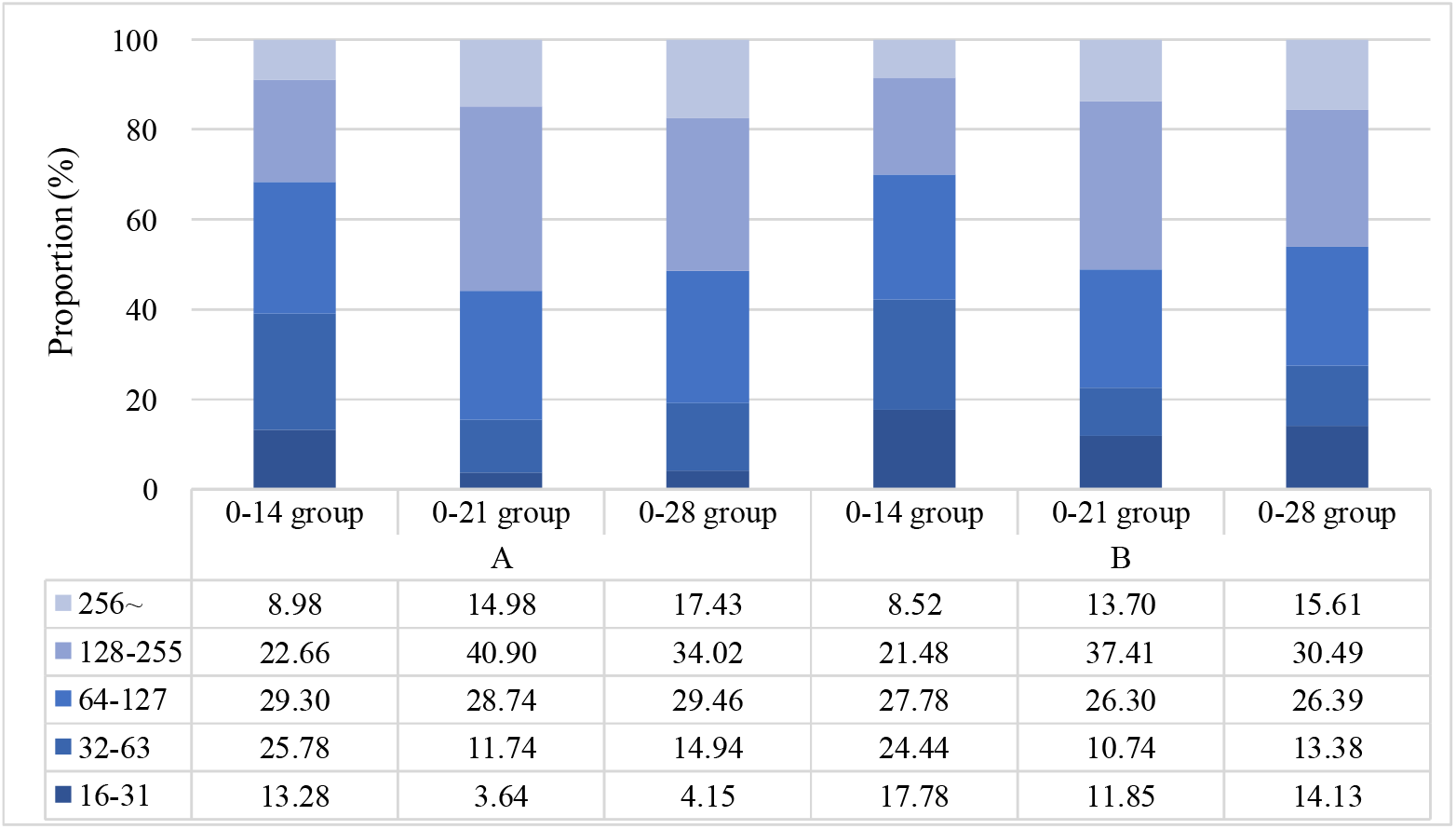
Distribution of SARS-CoV-2 Neutralizing Antibody. A: Per-protocol analysis, B: Modified intention-to-treat analysis. The table shows the percentages of SARS-CoV-2 neutralizing antibody in each group.

#### Stratified Analysis

In the further analysis stratified by age and gender, we observed the similar results that the GMT of SARS-CoV-2 neutralizing antibody and positive rates in the 0-21 and 0-28 groups were superior to the 0-14 group for the participants with different characteristic levels. (Supplement 2, Supplement 3). And the similar distribution results of SARS-CoV-2 neutralizing antibody were observed for the three groups. (Supplement 4).

### Influencing Factors of SARS-CoV-2 Neutralizing Antibody Immunization by Multinomial Logistic Regression

Multinomial logistic regression was performed to examine the influencing factors of SARS-CoV-2 neutralizing antibody immunization depending on their extent (GMT: 16-63=1, GMT: 64-127=2, GMT: 128∼=3, ref=1) and the results showed that only the vaccination regimen was associated with the antibody response. After adjusting for age, gender, BMI, marital status, education level, influenza vaccination history, smoking, drinking, and chronic diseases, participants who received 0-21 vaccination regimen was 2.51 times higher than the 0-14 vaccination group when GMT was 64-127 (95% *CI*: 1.52, 4.13), and it was 4.42-fold higher than that in 0-14 vaccination group at GMT ≥ 128 (95% *CI*: 2.77, 7.07). Participants in the 0-28 group had 2.00-fold odds when GMT was 64-127 (95% *CI*: 1.23, 3.25) than that in the 0-14 group and had 3.30-fold odds of GMT ≥ 128 (95% *CI*: 2.09, 5.22) (Table 3).

**Table 3.** Influencing Factors of SARS-CoV-2 Neutralizing Antibody Immunization by Multinomial Logistic Regression.

| Variables | OR (95% *CI*) | <sup>a</sup> OR (95% *CI*) * |
| --- | --- | --- |
| GMT 64-127 (ref: 16-63) |  |  |
| 0-14 group | 1.00 | 1.00 |
| 0-21 group | 2.49 (1.52, 4.09) | 2.51 (1.52, 4.13) |
| 0-28 group | 2.06 (1.28, 3.32) | 2.00 (1.23, 3.25) |
| GMT 128~ (ref: 16-63) |  |  |
| 0-14 group | 1.00 | 1.00 |
| 0-21 group | 4.48 (2.82, 7.13) | 4.42 (2.77, 7.07) |
| 0-28 group | 3.33 (2.13, 5.21 ) | 3.30 (2.09, 5.22) |

\* Adjusted by age, gender, BMI, marital status, education level, influenza vaccination history, smoking, drinking, and chronic diseases.

### Logistic Regression Analysis of Factors Influencing the SARS-CoV-2 Neutralizing Antibody Immunization

Multivariate analysis results showed that only vaccination regimen was associated with SARS-CoV-2 neutralizing antibody immunization, and there were no other factors were found to be associated with SARS-CoV-2 neutralizing antibody immunization. After adjusting for age, gender, BMI, marital status, education level, influenza vaccination history, smoking, drinking, and chronic diseases, the results showed that the participants in 0-21 and 0-28 groups was 4.10 (95% *CI*: 1.90, 8.84) and 3.59 (95% *CI:* 1.70, 7.57) times more likely to be positive (GMT ≥ 32) than those in 0-14 groups, respectively; the results showed that the participants in 0-21 and 0-28 groups showed higher positive rates than those in 0-14 group (GMT ≥ 64) (*OR*: 3.50, 95% *CI*: 2.27, 5.38; *OR*: 2.70, 95% *CI*: 1.78, 4.09). The similar results were found at GMT ≥ 128 or 256. (Table 4).

**Table 4.** Logistic regression analysis of Factors Influencing the SARS-CoV-2 Neutralizing Antibody Immunization.

| Variables | *OR (95% CI)* | *<sub>a</sub> OR (95% CI) \** |
| --- | --- | --- |
| GMT ≥ 32 | | |
| 0-14 group | 1.00 | 1.00 |
| 0-21 group | 4.05(1.90, 8.64) | 4.10(1.90, 8.84) |
| 0-28 group | 3.54(1.71, 7.33) | 3.59(1.70, 7.57) |
| GMT ≥ 64 | | |
| 0-14 group | 1.00 | 1.00 |
| 0-21 group | 3.53(2.30, 5.41) | 3.50(2.27, 5.38) |
| 0-28 group | 2.72(1.81, 4.09) | 2.70(1.78, 4.09) |
| GMT ≥ 128 | | |
| 0-14 group | 1.00 | 1.00 |
| 0-21 group | 2.74(1.90, 3.94) | 2.70(1.86, 3.90) |
| 0-28 group | 2.29(1.59, 3.30) | 2.33(1.60, 3.39) |
| GMT ≥ 256 | | |
| 0-14 group | 1.00 | 1.00 |
| 0-21 group | 1.79(1.03, 3.10) | 1.80(1.02, 3.19) |
| 0-28 group | 2.14(1.24, 3.68) | 2.25(1.28, 3.95) |

\* Adjusted by age, gender, BMI, marital status, education level, influenza vaccination history, smoking, drinking, and chronic diseases.

### Safety Outcomes

The overall incidence of adverse reactions was 11 (4.07%) of 270 in the days 0 and 14 vaccination cohort group, 13 (4.81%) of 270 in the days 0 and 21 vaccination cohort group, and 10 (3.72%) of 269 in the days 0 and 28 vaccination cohort group.

Solicited adverse reactions were reported by 26 (3.21%) within 7 days after injection, and 8 (0.99%) reported unsolicited adverse reactions within 28 days in the trials. No significant differences were found in the occurrence of solicited and unsolicited adverse reactions among the three groups. Pain, swelling, pruritus, diarrhea and fatigue within 7 days after vaccination were reported by 4 (1.48%), 2 (0.74%), 1 (0.37%), 0 (0.00%), and 1 (0.37%) subject in the 0-14 group, 7 (2.59%), 0 (0.00%), 2 (0.74%), 1 (0.37%), and 1 (0.37%) subject in the 0-21 group, and 2 (0.74%), 2 (0.74%), 0 (0.00%), 1 (0.37%), and 2 (0.74%) subject in the 0-28 vaccination group, respectively. Rash, cough and headache within 28 days after vaccination were reported by 1 (0.37%), 1 (0.37%), and 1 (0.37%) subject in the 0-14 group, 1 (0.37%), 1 (0.37%), and 0 (0.00%) subject in the 0-21 group, and 1 (0.37%), 1 (0.37%), and 1 (0.37%) subject in the 0-28 group, respectively. The reported adverse reactions did not differ significantly among the three study groups (*P* > 0.05).

None of the subjects reported serious adverse reactions or became SARS-CoV-2 infected during the follow-up period (Table 5).

**Table 5.** Summary of solicited and unsolicited adverse reactions occurred within 28 days during the study period.

| Adverse reaction * | 0-14 group (n=270) | 0-21 group (n=270) | 0-28 group(n=269) | *P* |
| --- | --- | --- | --- | --- |
| Solicited adverse reactions within 0-7days |  |  |  |  |
| Local reactions | 7(2.59) | 9(3.33) | 4(1.49) | 0.418 |
| Pain | 4(1.48) | 7(2.59) | 2(0.74) | 0.263 |
| Swelling | 2(0.74) | 0(0.00) | 2(0.74) | 0.479 |
| Pruritus | 1(0.37) | 2(0.74) | 0(0.00) | 0.777 |
| Systemic reactions | 1(0.37) | 2(0.74) | 3(1.12) | 0.545 |
| Diarrhea | 0(0.00) | 1(0.37) | 1(0.37) | 0.777 |
| Fatigue | 1(0.37) | 1(0.37) | 2(0.74) | 0.702 |
| Unsolicited adverse reactions within 8-28 days |  |  |  |  |
| Local reactions | 1(0.37) | 1(0.37) | 1(0.37) | 1.000 |
| Rash | 1(0.37) | 1(0.37) | 1(0.37) | 1.000 |
| Systemic reactions | 2(0.74) | 1(0.37) | 2(0.74) | 0.876 |
| Cough | 1(0.37) | 1(0.37) | 1(0.37) | 1.000 |
| Headache | 1(0.37) | 0(0.00) | 1(0.37) | 0.777 |

Results expressed as n (%); \* Adverse reaction data only list the occurrence of this symptom.

## Discussion

Given the COVID-19 pandemic continuing to unfold, a safe and effective vaccine is necessary to contain the global COVID-19 pandemic and prevent further illness and fatalities. Inactivated vaccines are generally safe and widely used for prevention of infectious diseases. On May 7, 2021, WHO approved the inactivated SARS-CoV-2 vaccine manufactured by the Beijing Institute of Biological Products/Sinopharm (China) in its Emergency Use Listing. This is the first randomized controlled trial for assessment of the immunogenicity and safety of an inactivated SARS-CoV-2 vaccine in high-risk occupational population.

Here, we explored the immunogenicity and safety of the three different SARS-CoV-2 inactivated vaccination schemes, and found that the GMT of neutralizing antibody were between 98.41 and 145.50, with the seroconversion rates (GMT ≥ 16) being 100% in the three groups. The current other clinical trials have also assessed safety and immunogenicity of SARS-CoV-2 vaccines and have seen comparable results. The existing inactivated virus vaccines have shown good antibody responses (79%-100%) and neutralizing antibody titers (18.9-282.7)^15-17, 22-24^. And a recent phase 3 clinical trials^20^ with two inactivated SARS-CoV-2 vaccines (WIV04, HB02) showed a vaccine efficacy of 64.0% (95% *CI*, 48.8%-74.7%) and 73.5% (95% *CI*, 60.6%-82.2%). In addition, the efficacy of other kinds of SARS-CoV-2 vaccines (such as mRNA vaccines, recombinant adenovirus-based vaccine, or chimpanzee adenoviral vector vaccine) in phase 3 clinical trials is approximately 70.4%-95.0%^18, 21, 25, 26^, of those, a study^21^ of an rAd26 and rAd5 vector-based heterologous prime-boost COVID-19 vaccine reported that the GMT of neutralizing antibody was 44.5 (95% *CI*, 31.8-62.2) and the seroconversion rate was 95.83% in the vaccine group. These studies indicated that the current SARS-CoV-2 vaccines have relatively good immune effect.

In our research, the SARS-CoV-2 neutralizing antibody titers of the 0-28, 0-21 groups were significantly greater than that of 0-14 group. Some trials have also designed to compare the immunogenicity of inactivated SARS-CoV-2 vaccination regimens in healthy adults^15-17, 22^. Xia^15, 16^ et al. in both phase 1 and 2 found that a longer interval (21 days and 28 days) produced higher antibody responses compared with a shorter interval schedule (14 days) (GMT: 282.7 [221.2-361.4]; 218.0 [181.8-261.3] vs. 169.5 [132.2-217.1]). Similarly, a study by Zhang^17^ et al. showed that after two doses of inactivated SARS-CoV-2 vaccine, the SARS-CoV-2 neutralizing antibody titers and seroconversion rates induced by 0-28 vaccination schedule (97%, 44.1 [37.2-52.2] in the 3 µg group; 100%, 65.4 [56.4-75.9] in the 6 µg group) were higher than those induced by 0-14 vaccination schedule (94%, 23.8 [20.5-27.7] in the 3 µg group; 99%, 30.1 [26.1-34.7] in the 6 µg group). Pan^22^ et al. also found that the 0-28 regimen of inactivated SARS-CoV-2 vaccine induced higher SARS-CoV-2 neutralizing antibody titers and seroconversion rates (5 µg: 99.0%, 131.7[109.3-158.6]; 10 µg: 100.0%, 110.7 [94.7-129.4]) compared with the 0-14 regimen (5 µg: 98.0%, 37.2 [29.5-46.9]; 10 µg: 96.0%, 44.5 [35.5-55.7]). And at 28 days after the second dose, the GMT induced by the 0-28 regimen are 1.6 to 3.5 times higher than that in the 0-14 regimen (*P* < 0.0005)^22^. All of above studies indicated that longer interval schedule (0-21 regimen or 0-28 regimen) of the inactivated SARS-CoV-2 vaccines may induce a better immunogenicity, which was certified in the recent studies of phase 3 clinical trial^18, 20, 21^. A study^20^ using 2 inactivated SARS-CoV-2 vaccines with 21-day intervals in several countries showed 99.3% (WIV04) and 100.0% (HB02) seroconversion rates, and a vaccine efficacy of 64.0% (95% *CI*, 48.8%-74.7%) and 73.5% (95% *CI*, 60.6%-82.2%) against COVID-19. Similarly, other mRNA vaccine and vector-based vaccine in phase 3 trial^18, 21^ also elicited a better immunogenicity at an interval of 0-21 days, with the presence of neutralizing antibody in more than 90% of the participants. In our study, the neutralizing antibody titer in 0-21 group was numerically higher than that of 0-28 group. The current studies have indicated that an interval of both 0-21 and 0-28 days elicited significantly higher SARS-CoV-2 neutralizing antibody levels than 0-14 days interval, with a numerically higher neutralizing antibody level in 0-21 days interval, suggesting that the 0-21 days interval schedule of the SARS-CoV-2 vaccines may be a better regimen.

The incidence of adverse reactions in the 0-14, 0-21 and 0-28 groups were similarly low. Moreover, we did not find severe adverse reaction, with the most common symptom being injection-site pain, indicating no safety concerns. The overall incidence of adverse events after vaccination was 3.72-4.81% in our vaccine-treated groups, which is noticeably lower than that of other SARS-CoV-2 vaccine platform candidates such as viral-vectored vaccines, DNA or RNA vaccines^26-31^. The safety profile of this vaccine in our study is also lower than that of other inactivated SARS-CoV-2 vaccines^15, 17^, which may be related to different population characteristics, and minor adverse reactions that are not reported.

Our study had several limitations. First, we only reported immune response data for the high-risk occupational population aged 18 to 59 years. Further studies are required to assess the immunogenicity of inactivated SARS-CoV-2 vaccine in various populations, including general population, older people, children and adolescents. Second, data on long-term immunogenicity is not yet available, and the ongoing trial will provide more information. Third, cellular immunity and immune memory were not measured in the current study which need to be further studied.

In summary, a two-dose of inactivated SARS-CoV-2 vaccine at 0-21 days and 0-28 days regimens significantly improved SARS-CoV-2 neutralizing antibody level compared to the 0-14 days regimen in high-risk occupational population. And the neutralizing antibody titer in 0-21 group was numerically higher than that in 0-28 group.

## Data Availability

The full study protocol and the datasets, which includes all data fields reported in this study, are available, following manuscript publication, upon request from the corresponding author (Professor Suping Wang), following the provision of ethics approval.

## Acknowledgements

This research was supported by the COVID-19 Project of Shanxi Provincial Finance, and the Project of Shanxi Provincial Key Laboratory for major infectious disease response. Thanks for all participants taking part in this research. We gratefully acknowledge the contribution from our colleagues and students, and staff members of the Shanxi Provincial Center for Disease Control and Outpatient Department of Shanxi Aviation Industry Group Co. LTD.

## Author contributions

Yongliang Feng: conceptualization, conceived and developed the overall study design, reviewed and interpreted data, investigated and performed experiments, and drafted the manuscript Jing Chen, Tian Yao: conceptualization, conceived and designed the study, analyzed the data, reviewed and interpreted data, investigated and performed experiments. Yue Chang: analyzed the data, reviewed and interpreted data, investigated and performed experiments. Xiaoqing Li, Rongqin Xing, Hong Li, Ruixue Xie, Xiaohong Zhang, Zhiyun Wei, Shengcai Mu, and Ling Liu: investigated and performed experiments. Lizhong Feng* & Suping Wang*: conceptualization, conceived and designed the study, reviewed and interpreted data, writing-review & editing.

