## Supplementary material for "Safety and Immunogenicity of Inactivated SARS-CoV-2 Vaccine in High-Risk Occupational Population: a randomized, parallel, controlled clinical trial": Supplement 1.docx

**Supplement 1.** Demographic and Behavioral Characteristics of High-risk Occupational Population 28 Days after the Whole Course of Vaccination

| Characteristics | Total (n=744) | 0-14 group (n=256) | 0-21 group (n=247) | 0-28 group(n=241) | *P* |
| --- | --- | --- | --- | --- | --- |
| Gender |  |  |  |  | 0.493 |
| Male | 538(72.31) | 179 (69.92) | 179(72.47) | 180(74.69) |  |
| Female | 206(27.69) | 77(30.08) | 68(27.53) | 61(25.31) |  |
| Age(year) |  |  |  |  | 0.409 |
| <40 | 427(57.39) | 139(54.30) | 143(57.89) | 145(60.17) |  |
| 40~ | 317(42.61) | 117(45.70) | 104(42.11) | 96(39.83) |  |
| Education level |  |  |  |  | 0.222 |
| Junior high school or lower | 70(9.41) | 31(12.11) | 23(9.32) | 16(6.64) |  |
| Senior high school | 33(4.44) | 10(3.91) | 9(3.64) | 14(5.81) |  |
| College or higher | 641(86.15) | 215(83.98) | 215(87.04) | 211(87.55) |  |
| Ethnicity |  |  |  |  | 0.481^a^ |
| Han ethnicity | 733(98.52) | 254 (99.22) | 242(97.98) | 237(98.34) |  |
| other | 11(1.48) | 2(0.78) | 5(2.02) | 4(1.66) |  |
| Marital status |  |  |  |  | 0.161 |
| Married | 573(77.02) | 205(80.08) | 179(72.47) | 189(78.43) |  |
| Unmarried | 150(20.16) | 45(17.58) | 62(25.10) | 43(17.84) |  |
| Divorced or Widowed | 21(2.82) | 6(2.34) | 6(2.43) | 9(3.73) |  |
| BMI (kg/m^2^) |  |  |  |  | 0.858 |
| <18.5 | 19(2.55) | 8(3.13) | 6(2.43) | 5(2.07) |  |
| 18.5~ | 308(41.40) | 107(41.80) | 106(42.91) | 95(39.42) |  |
| 24~ | 417(56.05) | 141(55.07) | 135(54.66) | 141(58.51) |  |
| Influenza vaccination history |  |  |  |  | 0.930 |
| No | 497(66.80) | 169(66.02) | 167(67.61) | 161(66.80) |  |
| Yes | 247(33.20) | 87(33.98) | 80(32.39) | 80(33.20) |  |
| Occupation |  |  |  |  | 0.932 |
| Public security officers | 362(48.66) | 125(48.83) | 122(49.39) | 115(47.72) |  |
| Airport ground staff | 382(51.34) | 131(51.17) | 125(50.61) | 126(52.28) |  |
| Smoking |  |  |  |  | 0.227 |
| No | 501(67.34) | 182(71.09) | 165(66.80) | 154(63.90) |  |
| Yes | 243(32.66) | 74(28.91) | 82(33.20) | 87(36.10) |  |
| Drinking |  |  |  |  | 0.960 |
| No | 568(76.34) | 197(76.95) | 188(76.11) | 183(75.93) |  |
| Yes | 176(23.66) | 59(23.05) | 59(23.89) | 58(24.07) |  |
| Chronic diseases |  |  |  |  | 0.954 |
| No | 689(92.61) | 238(92.97) | 229(92.71) | 222(92.12) |  |
| Yes | 55(7.39) | 18(7.03) | 18(7.29) | 19(7.88) |  |

Results expressed as n (%); ^a^ Fisher’s exact test.
