## Supplementary material for "Safety and Immunogenicity of Inactivated SARS-CoV-2 Vaccine in High-Risk Occupational Population: a randomized, parallel, controlled clinical trial": Supplement 2.docx

**Supplement 2.** SARS-CoV-2 Neutralizing Antibody Immunization in the Three Groups 28 Days after the Whole Course of Vaccination Stratified by Age and Gender

a. SARS-CoV-2 neutralizing antibody immunization by age

| SARS-CoV-2 neutralizing antibody | age<40 | | |  | age≥40 | | |
| --- | --- | --- | --- | --- | --- | --- | --- |
|  | 0-14 group  (n=139) | 0-21 group  (n=143) | 0-28 group  (n=145) |  | 0-14 group  (n=117) | 0-21 group  (n=104) | 0-28 group  (n=96) |
| GMT ≥ 32 | |  |  |  |  |  |  |
| No n(%) | 25(17.99) | 5(3.50) | 7(4.83) |  | 9(7.69) | 4(3.85) | 3(3.13) |
| Yes n(%) | 114(82.01) ^a^ | 138(96.50) ^b^ | 138(95.17) |  | 108(92.31) | 100(96.15) | 93(96.87) |
| GMT ≥ 64 | |  |  |  |  |  |  |
| No n(%) | 54(38.85) | 19(13.29) | 35(24.14) |  | 46(39.32) | 19(18.27) | 11(11.46) |
| Yes n(%) | 85(61.15) ^a^ | 124(86.71) ^b^ | 110(75.86) ^a^ |  | 71(60.68) ^a^ | 85(81.73) ^b^ | 85(88.54) ^b^ |
| GMT ≥ 128 | |  |  |  |  |  |  |
| No n(%) | 96(69.06) | 65(45.45) | 74(51.03) |  | 79(67.52) | 44(42.31) | 43(44.79) |
| Yes n(%) | 43(30.94) ^a^ | 78(54.55) ^b^ | 71(48.97) ^b^ |  | 38(32.48) ^a^ | 60(57.69) ^b^ | 53(55.21) ^b^ |
| GMT ≥ 256 | |  |  |  |  |  |  |
| No n(%) | 122(87.77) | 124(86.71) | 121(83.45) |  | 111(94.87) | 86(82.69) | 78(81.25) |
| Yes n(%) | 17(12.23) | 19(13.29) | 24(16.55) |  | 6(5.13) ^a^ | 18(17.31) ^b^ | 18(18.75) ^b^ |

b. SARS-CoV-2 neutralizing antibody immunization by gender

| SARS-CoV-2 neutralizing antibody | male | | |  | female | | |
| --- | --- | --- | --- | --- | --- | --- | --- |
|  | 0-14 group  (n=179) | 0-21 group  (n=179) | 0-28 group  (n=180) |  | 0-14 group  (n=77) | 0-21 group  (n=68) | 0-28 group  (n=61) |
| GMT ≥ 32 | |  |  |  |  |  |  |
| No n(%) | 23(12.85) | 5(2.79) | 6(3.33) |  | 11(14.29) | 4(5.88) | 4(6.56) |
| Yes n(%) | 156(87.15) ^a^ | 174(97.21) ^b^ | 174(96.67) |  | 66(85.71) | 64(94.12) | 57(93.44) |
| GMT ≥ 64 | |  |  |  |  |  |  |
| No n(%) | 70(39.11) | 25(13.97) | 35(19.44) |  | 30(38.96) | 13(19.12) | 11(18.03) |
| Yes n(%) | 109(60.89) ^a^ | 154(86.03) ^b^ | 145(80.56) |  | 47(61.04) ^a^ | 55(80.88) | 50(81.97) ^b^ |
| GMT ≥ 128 | |  |  |  |  |  |  |
| No n(%) | 123(68.72) | 76(42.46) | 88(48.89) |  | 52(67.53) | 33(48.53) | 29(47.54) |
| Yes n(%) | 56(31.28) ^a^ | 103(57.54) ^b^ | 92(51.11) ^b^ |  | 25(32.47) ^a^ | 35(51.47) | 32(52.46) ^b^ |
| GMT ≥ 256 | |  |  |  |  |  |  |
| No n(%) | 163(91.06) | 153(85.47) | 151(83.89) |  | 70(90.91) | 57(83.82) | 48(78.69) |
| Yes n(%) | 16(8.94) | 26(14.53) | 29(16.11) |  | 7(9.09) | 11(16.18) | 13(21.31) |

^a b^ There was significant difference with the different letters.
