## Supplementary material for "Safety and Immunogenicity of Inactivated SARS-CoV-2 Vaccine in High-Risk Occupational Population: a randomized, parallel, controlled clinical trial": Supplement 3.docx

**Supplement 3.** GMT of SARS-CoV-2 Neutralizing Antibody in the Three Groups 28 Days after the Whole Course of Vaccination Stratified by Age and Gender

GMT

|  | GMT |
| --- | --- |
| age<40 0-14 group | 101.2 |
| age<40 0-21 group | 131.9 |
| age<40 0-28 group | 141.5 |
| age≥40 0-14 group | 95.11 |
| age≥40 0-21 group | 137.8 |
| age≥40 0-28 group | 151.5 |

GMT

|  | GMT |
| --- | --- |
| male 0-14 group | 96.94 |
| male 0-21 group | 138.8 |
| male 0-28 group | 144 |
| female 0-14 group | 101.8 |
| female 0-21 group | 122.7 |
| female 0-28 group | 149.6 |

a. GMT of SARS-CoV-2 neutralizing antibody by age b. GMT of SARS-CoV-2 neutralizing antibody by gender
