## Supplementary material for "Safety and Immunogenicity of Inactivated SARS-CoV-2 Vaccine in High-Risk Occupational Population: a randomized, parallel, controlled clinical trial": Supplement 4.docx

**Supplement 4.** Distribution of SARS-CoV-2 Neutralizing Antibody in the Three Groups 28 Days after the Whole Course of Vaccination Stratified by Age and Gender

a

Proportion (%)

|  | 16~ | 32~ | 64~ | 128~ | 256~ |
| --- | --- | --- | --- | --- | --- |
| age<40 0-14 group | 17.99 | 20.86 | 30.22 | 18.71 | 12.22 |
| age<40 0-21 group | 3.49 | 9.79 | 32.17 | 41.26 | 13.29 |
| age<40 0-28 group | 4.83 | 19.31 | 26.9 | 32.41 | 16.55 |
| age≥40 0-14 group | 7.69 | 31.62 | 28.21 | 27.35 | 5.13 |
| age≥40 0-21 group | 3.85 | 14.42 | 24.04 | 40.38 | 17.31 |
| age≥40 0-28 group | 3.13 | 8.33 | 33.33 | 36.46 | 18.75 |

b

Proportion (%)

|  | 16~ | 32~ | 64~ | 128~ | 256~ |
| --- | --- | --- | --- | --- | --- |
| male 0-14 group | 12.85 | 26.26 | 29.61 | 22.35 | 8.93 |
| male 0-21 group | 2.79 | 11.17 | 28.49 | 43.02 | 14.53 |
| male 0-28 group | 3.33 | 16.11 | 29.44 | 35.01 | 16.11 |
| female 0-14 group | 14.29 | 24.68 | 28.57 | 23.38 | 9.08 |
| female 0-21 group | 5.88 | 13.24 | 29.41 | 35.29 | 16.18 |
| female 0-28 group | 6.55 | 11.48 | 29.51 | 31.15 | 21.31 |

a. Distribution of SARS-CoV-2 neutralizing antibody by age b. Distribution of SARS-CoV-2 neutralizing antibody by gender
